# Disclosing of participation in an HIV vaccine trial in southwestern Uganda – the effect on participant engagement

**DOI:** 10.1101/2025.09.11.25334968

**Authors:** Sarah Nakamanya, Rachel Kawuma, Denis Kibuuka, Georgina Nabaggala, Sylvia Kusemererwa, Eugene Ruzagira, Janet Seeley, the PrEPVacc Study Group

## Abstract

Developing a safe, effective HIV vaccine remains important for the control of the epidemic. However, the development and testing of such a vaccine faces a range of social and behavioural challenges including potential stigmatisation, leading to non-disclosure of trial participation. We explored disclosure of participation in an HIV PrEP and vaccine trial to understand its effect on participant engagement.

Between 22 September 2021 and 8 August 2023, 5% (30) of individuals in an HIV vaccine trial (PrEPVacc trial [NCT04066881]) were purposively selected for repeat in-depth interviews at 2, 6 and 12 months of participation in the trial. Forty-five other individuals took part in six focus group discussions, divided equally by gender, with each group comprising 6 to 10 participants. Experiences with vaccination, motivation to participate, and disclosure of participation were explored. Data were analysed thematically using a manual framework analysis approach.

Disclosure was reported to enhance participation. Participants disclosed to friends, sexual partners, family members, and employers. Reasons for disclosure included a desire to participate openly and comfortably, to gain others’ support including financial and physical, and to motivate others. Men and younger participants disclosed more readily than women and older participants. Concerns like fear of being suspected of infidelity or having HIV, blame, and relationship breakages hampered disclosure. A lack of confidence in explaining what the study was about prevented disclosure, as a failure to explain attracted negative reactions including disapproval. As participants gained confidence in the study, approval and support from others improved. Participants who disclosed their participation tended to adhere to study requirements better and encountered less discomfort about taking part, or fear of potential social harm.

Successful HIV vaccine trial participation calls for proactive public engagement and awareness campaigns. Disclosure by study participants to significant others about taking part could increase participation, strengthen retention and adherence.

## Introduction

HIV remains a global problem, as 1.3 million new infections were recorded in 2023, of which 450,000 occurred in Eastern and Southern Africa(1). In 2023, an estimated 20,000 Ugandans died of HIV-related illnesses, and 730 new infections were registered every week, with a total of 38,000 new infections registered that year (2).

Finding an effective HIV vaccine would be an important contribution to ending the AIDS epidemic by 2030 (3, 4). Efforts to find a vaccine are ongoing. In 2009, an HIV vaccine trial in Thailand demonstrated modest efficacy at 31.2% (5) while other trials ended prematurely including the HVTN 702, which was stopped in January 2020 for not demonstrating any efficacy (4, 6–9).

However, HIV vaccine trials have faced various barriers including participants expressing doubt about trial participation risks, such as acquiring HIV infection (10, 11). Vaccine induced sero-positivity (VISP), the detection of vaccine-induced antibodies during HIV testing (12), continues to present challenges for participants affected, impacting their relationships, with possible consequences for travel and employment. Other concerns include limited knowledge of research trials, and fear of being assumed to be HIV-positive because of trial participation (13) as well as vaccine side effects (14). Concerns linked to HIV-related stigma, are significant barriers to participating in HIV clinical trials (15, 16). Stigma and the fear of being stigmatised not only discourage participation in HIV-related interventions and research but can also have an impact on the willingness to disclose involvement in a trial. A clear understanding of the factors around disclosure and the impact on participant engagement is important.

In this paper we examine how participants in Uganda taking part in an HIV PrEP and vaccine trial (the PrEPVacc trial) understood ‘HIV vaccines’ and how disclosing their taking part influenced their participation in the trial.

We define participant engagement as actively involving and keeping study participants informed, interested, and engaged (active) throughout the entire research process, from recruitment to post-study activities.

### The Disclosure Processes Model

To help us understand how trial participants navigated through disclosure, including the decision to disclose their involvement and how this affected their participation, we employed the Disclosure Processes Model (DPM). The model provides a theoretical framework to understand why and when interpersonal, verbal self-disclosure is beneficial for individuals who live with concealable stigmatised identities, such as HIV (17). According to the DPM, disclosure is influenced by individual motivations, anticipated outcomes, and the social context in which disclosure occurs.

The DPM suggests that a person’s goals, whether to seek positive outcomes or avoid negative ones, affect how disclosure impacts personal, relational, and social outcomes. These effects happen through three key processes: (1) alleviating inhibition (reducing stress or hesitation), (2) gaining social support, and (3) influencing how others perceive and respond to the disclosure (17).

Understanding these dynamics through the DPM lens can help researchers develop strategies to support participants in making informed disclosure decisions that promote sustained engagement in HIV vaccine and other clinical trials.

## Methods

### Study design and setting

This qualitative study was part of the PrEPVacc trial (NCT04066881), a Phase IIb three-arm, two-stage HIV prophylactic vaccine trial with a second randomisation to compare TAF/FTC to TDF/FTC as pre-exposure prophylaxis. It was a multi-site trial conducted in South Africa, Tanzania, and in Uganda at the Medical Research Council /Uganda Virus Research Institute & London School of Hygiene and Tropical Medicine (MRC/UVRI & LSHTM), Uganda Research Unit from 2018 to 2024. Men and women aged 18-40 years who were at risk of HIV acquisition were enrolled in the trial, initially through an HIV preparedness registration cohort (18), which preceded enrolment in the main trial, with additional enrolment from the same communities before the main trial began. In this paper we focus on participants from the Uganda site.

Before enrolment, participants received basic information about the planned trial, including that two HIV vaccine combinations which were to be tested against a placebo to find out if they were efficacious. They got information on the different study procedures, including the qualitative study. Potential participants were identified from fishing communities around Lake Victoria and trading towns along the Trans-African highway in Masaka district in southwestern Uganda. Fisherfolk, female sex workers, and other groups at higher risk for HIV acquisition were recruited for the trial. Participants were randomly assigned in a 1:1:1 ratio to either of two experimental HIV vaccines [DNA-HIV-PT123 and AIDSVAX® B/E (at week 0, 4, 24, 48) and DNA-HIV-PT123 and CN54gp140+MPLA-L (at week 0, 4), then MVA-CMDR and CN54gp140+MPLA-L (week 24, 48)], or placebo. The second randomization, at a ratio of 1:1 compared two different types of oral PrEP regimens; Tenofovir Alafenamide/emtricitabine (TAF/FTC) and Tenofovir disoproxil fumarate/emtricitabine (TDF/FTC). Participants continued to receive study PrEP up to two weeks after the third injection, after which access to PrEP was available through local clinics, for participants who wished to continue with PrEP. The Uganda site enrolled a total of 512 participants (87% women) in the trial.

### Sampling procedure

Between 22 September 2021 and 8 August 2023, about 5% (30) of trial participants were selected to take part in repeat in-depth interviews at 2, 6 and 12 months during the trial. They were purposively selected to represent patterns of HIV risk and adherence from quantitative trial data. Sample characteristics included the different age groups, gender, socio-economic backgrounds or work type and PrEP regimen. In terms of PrEP use, we included those who adhered well (someone with consistent results between self-report and urine over a period of 3 visits) the not so good adherers (some inconsistent results) and poor adherers (those with results showing that PrEP had not been taken) in the sample.

The data manager at the site generated a list based on the above characteristics. The social scientists, undertaking the qualitative study, worked closely with the counsellors and identified these participants at the week 4 visit and scheduled an appointment to see the participant again for the baseline in-depth interview when they came for their week 8 visit (month 2). In cases where this was not possible, a call was made to the participant to schedule an appointment for the in-depth interview (IDI).

A further sample of 45 individuals was selected to take part in focus group discussions (FGDs). Groups were divided equally by gender and comprised 6 to 10 participants and were conducted with other study participants not participating in the IDIs. FGDs followed the same sampling strategy as the in-depth interviews whereby characteristics such as age, gender, socio-economic background and PrEP regimen were considered. Discussants and interviewees represented good adherers, poor adherers, and PrEP refusers.

### Data collection

Two (male and female) experienced research assistants interviewed participants, and these were matched by gender. A generic interview guide, translated into the local language (Luganda) was used at the different phases of the study. Baseline interviews were conducted after two months of participation in the trial. These collected background information of participants as well as experiences of using PrEP and vaccination. The second interview was conducted after 6 months (after 26 weeks) of participation in the trial, after participants had stopped taking study PrEP, to establish whether they continued with PrEP at accepted referral to other PrEP facilities, support or barriers of adherence in the study, as well as experiences with the vaccine and trial in general. Information on knowledge of HIV prevention methods, views and fears regarding participation in vaccine trials, disclosure of their participation in the study and the reasons for disclosure/non-disclosure were also captured. The third interview was at month 12, (after one year of participation), we continued to explore the longer-term experiences that participants had during the trial.

FGDs were conducted at week 30, after participants had stopped the trial PrEP, and at week 48. These were conducted with participants who adhered well, those who did not adhere very well to PrEP, and another set with PrEP refusers to explore barriers and facilitators of adherence to PrEP and experiences in the trial in general. A total of 6 FGDs were conducted, facilitated by an experienced moderator, who guided the discussions, and a notetaker who recorded the conversations and observations.

Experiences with vaccination, motivation to participate and disclosure of participation were explored. Other issues discussed included HIV risk, knowledge and views of vaccines and attitudes towards participating in vaccine studies. IDIs and FGDs were conducted face-to-face in a convenient and private place at the clinic. Each IDI lasted for an average of one hour while FGDs lasted for an average of 90 minutes. All IDIs and FGDs were audio-recorded. Debriefing meetings with the research assistants and the lead social scientist were held periodically to discuss the interviews and ensure completeness of the data. Any identified gaps were addressed during the subsequent interviews.

### Data management and analysis

Audio recordings from IDIs and FGDs were transcribed verbatim and translated to English. Typed data transcripts were anonymized using identification numbers, stored on password-protected computers, and backed up on a secure server with restricted access to the study team.

Data analysis followed a framework analysis approach which involved identifying both inductive (new from the data) and deductive (from the topic guide) themes, indexing or coding, and charting, copying and pasting data according to relevant thematic areas. Mapping, which is the visual display of data, was done to allow researchers to identify patterns, associations, and concepts.

Analysis was done by the two interviewers with support from the lead social scientist. The initial stages involved interviewers independently reading two similar interview transcripts and identifying codes relevant to the research question. These were jointly agreed upon by the team and grouped together to form broader themes. Four dominant themes were identified for the analysis for this paper, informed by the ‘key processes’ of the DPM, namely: ‘Knowledge of HIV trial vaccines’, ‘Disclosure of participation’, ‘Benefits’ and ‘Concerns’ around disclosing. A final coding framework was agreed upon, and data were manually coded onto a matrix including illustrative quotes from the interviews under relevant themes, which we present in this paper.

### Ethical considerations

Approval for the PrEPVacc trial, which this qualitative study is part of, was obtained from the Uganda Virus Research Institute Research Ethics Committee (UVRI REC) (REF: GC/127/19/11/739), the Ugandan National Council for Science and Technology (UNCST), (REF: HS2717) and London School and Hygiene and Tropical Medicine (LSHTM) Ethics committee (REF:21317). Written informed consent was obtained from all participants prior to their enrolment in the main trial. A separate consent process was conducted for participation in the qualitative component, which included audio-recording of the interviews. Identified social harms were addressed either directly at the research site or through referrals to appropriate service providers for further support.

## Results

A total of 75 men and women participated in IDIs and FGDs. Of the 30 IDI participants, three did not complete month 12 interview due to relocation. More than two thirds of the participants were below the age of 30 years and were not married or cohabiting with a partner.

**Table 1:**
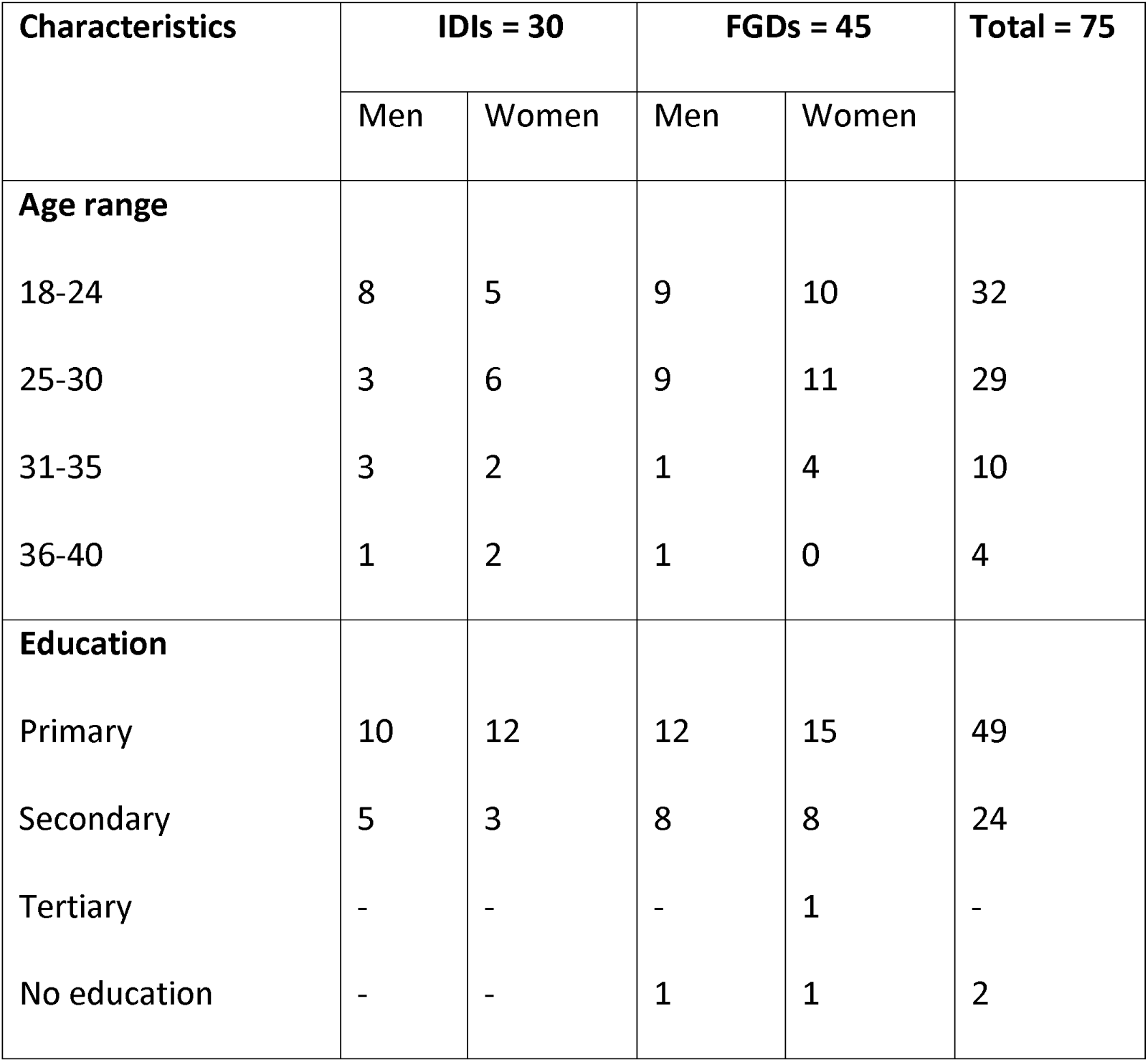
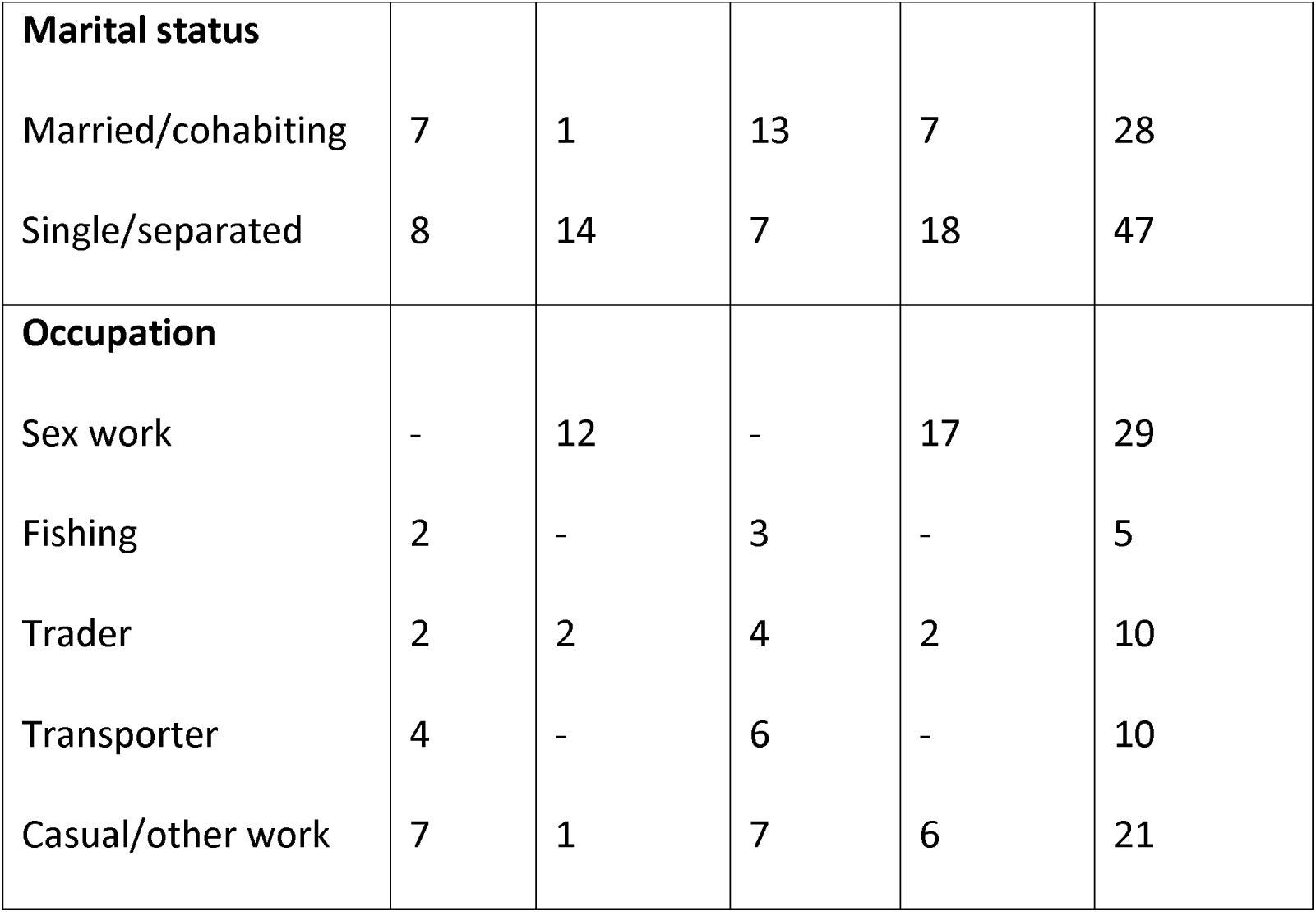
Socio-demographic information of participants.

| Characteristics | IDIs = 30 |  | FGDs = 45 |  | Total = 75 |
| --- | --- | --- | --- | --- | --- |
|  | Men | Women | Men | Women |  |
| **Age range** |  |  |  |  |  |
| 18-24 | 8 | 5 | 9 | 10 | 32 |
| 25-30 | 3 | 6 | 9 | 11 | 29 |
| 31-35 | 3 | 2 | 1 | 4 | 10 |
| 36-40 | 1 | 2 | 1 | 0 | 4 |
| **Education** |  |  |  |  |  |
| Primary | 10 | 12 | 12 | 15 | 49 |
| Secondary | 5 | 3 | 8 | 8 | 24 |
| Tertiary | - | - | - | 1 | - |
| No education | - | - | 1 | 1 | 2 |
| **Marital status** |  |  |  |  |  |
| Married/cohabiting | 7 | 1 | 13 | 7 | 28 |
| Single/separated | 8 | 14 | 7 | 18 | 47 |
| **Occupation** |  |  |  |  |  |
| Sex work | - | 12 | - | 17 | 29 |
| Fishing | 2 | - | 3 | - | 5 |
| Trader | 2 | 2 | 4 | 2 | 10 |
| Transporter | 4 | - | 6 | - | 10 |
| Casual/other work | 7 | 1 | 7 | 6 | 21 |

Participants’ narratives indicated that at baseline, a large number of them had not disclosed their participation in the trial but at the time of follow-up, most had disclosed to at least one person. They noted that disclosing that they were taking part in the trial had improved their participation because they were able to do it openly, without having to hide, and had gained the support and approval of the people they disclosed to. Participants also noted that disclosure was mostly to people they thought mattered to them or were significant in their lives.

### Knowledge about HIV trial vaccines

We explored what knowledge or information the participants or their communities had about HIV trial vaccines, and we found that none of the participants had any prior knowledge but acknowledged hearing about HIV vaccines for the first time after joining the registration cohort or the trial. Even when they discussed further about an HIV trial vaccine within their communities after hearing about it, other people in their communities denied that there could be an HIV vaccine.

“…they cannot understand even if I explain to them. I will explain it to them, until they understand it.” (male aged between 18 and 24 years)

However, for others the idea of an HIV vaccine seemed exciting, with participants claiming it would save them from acquiring or the worry of acquiring HIV since they had sex with many partners whose HIV status was unknown to the participants.

“…I want to remain healthy, since I am in multiple relationships and my husband too is in multiple relationships, so, the risk of getting HIV is high.…the reason is mostly to stay safe from HIV.” (female aged between 18 and 24 years)

However, given their prior knowledge of how other vaccines worked, they wondered what the HIV vaccine would be and whether they were going to be injected with the HIV virus itself as they believed was the case with other vaccines.

“Others say they are going to inject us with HIV and after that give us the drug. Is it true that they will first inject us with the virus?” (FGD, women)

Even with the study information that participants got, their narratives at baseline showed they had remembered a limited amount of information regarding HIV trial vaccines and had many questions about how these worked. This was aggravated by the wrong information within communities which sometimes distorted the study information given to participants and increased their fears. However, over time participants’ understanding of the trial and how the HIV trial vaccine worked improved.

“We sometimes try to encourage people to join but the number of questions they ask make you regret telling them. They ask you; ‘what is being tried? Why are they injecting people with that drug? Should it be tested on people or animals?’ That is why I ignore them”. (female aged between 25 and 30 years)

In that regard, participants attributed trial non-participation to the limited information, and they suggested that there was need to sensitize the communities and provide them with information on HIV vaccines and how they worked, to help promote use of these vaccines. They believed that communities needed to know how vaccines are manufactured and how trials are conducted as this would give them confidence of the vaccines’ safety.

Individuals in in-depth interviews noted that people who have participated in vaccine trials before would be the best to testify about vaccines to the rest of the population. According to them, these would be easily trusted, and relaying vaccine information would be very easy after conducting successful trials, as former trial participants would be the living examples and would just need to share their experiences.

“People will accept the vaccine if they get information from the people who have participated in a trial(s) before…such people can explain to them, and they can easily trust and believe”. (male aged between 25 and 30 years)

Such claims became a reality when individuals who had initially feared to take up the vaccines wanted to get vaccinated after seeing that others who had participated and completed their vaccination schedules did not have any health complications.

Thus, an understanding of the trial alleviated stress about being associated with the trial, and influenced whether participants disclosed their participation, which in turn shaped their engagement and participation.

### Disclosure of participation

Whereas participants were encouraged to disclose their trial participation to their primary health care providers (to avoid harmful drug interactions or for proper interpretation of symptoms and laboratory results), and to people they thought mattered in their lives and could influence their participation, disclosing was not mandated. The purpose of encouraging disclosure was to ensure a meaningful and successful participation in the trial.

While many individuals involved in the trial reported disclosing their participation to others, this varied in terms of whom they disclosed to and how much information they gave, reasons for disclosing, the approach or manner in which participation was revealed and the timing, and what facilitated their disclosure. The decision to disclose to someone and the amount of information given was partly determined by the closeness of the relationship. Generally, disclosure was to partners, peers/friends, immediate family (parents, siblings, children), employers, workmates, fellow study participants, and occasionally to other members of the community.

### Forms of Disclosure

Full disclosure - In some cases, participants disclosed all the details regarding the study they were involved in, including sharing the study information sheets. Detailed information about their participation was usually given to peers/friends and some sexual partners. Full or detailed disclosure usually happened where participants felt close and comfortable with the person being disclosed to, with no fear of being asked many questions or being judged. Other cases of full disclosure were when participants anticipated support from the person or because they wanted to encourage that person to join the trial.

“The people I mobilize to join know about it because I have to fully explain it to them in order to convince them.” (male aged between 31 and 35 years)

Partial disclosure - In other cases, disclosure was partial, and participants only shared specific aspects of their involvement or of the trial depending on the type of relationship to the individual they were disclosing to. For example, employers and other family members were usually given partial information. For example, a man aged between 36 and 40 years was asked if he disclosed to his current employer about his participation in the trial, he said; “I disclosed to them but did not give them the details. I just told them that I am participating in some study at the hospital, but I did not provide details”. Such disclosure was usually necessitated by the need to explain requests for time off to facilitate attendance at appointments for the trial.

A man aged between 18 and 24 years gave his parent partial information in case the clinic team contacted him via his parent: “I disclosed my participation in the trial to my current partner and to my parent although I did not explain the details to my parent. I only disclosed to them just in case I do not have a phone, and they want to contact me. I provided my parent’s phone number as the next of kin in case they (study staff) wanted to contact me”.

In their narratives, participants explained how they did not disclose to everyone at work or in their homes, if they did not need them to know for practical reasons, as in the cases above, because as a man aged between 18 and 24 years explained, he feared the information could be distorted:

’’The other people like the ones I work with, I cannot tell them because it would take me ages to explain to them so that they understand. They will keep on twisting the information from one person to the other. They will end up saying that I have HIV and I take ARVs. That is why I do not even labour to explain to anyone’’.

Being the household head and or provider, male participants disclosed their participation to their partners more easily than women. According to them, they were not answerable to anyone and hence no one would influence their decisions within their relationships.

“My partner would have to agree since she cannot influence my decisions or even influence me to change my mind”. (male aged between 18 and 24 years)

“I am an adult, and I know what I want, and it was voluntary; I keep my pills in a place visible to everyone. Even if you were my friend, leaving me because you saw me taking PrEP confirms that you were not a true friend” (male aged between 25 and 30 years).

Almost all men in in-depth interviews had disclosed to at least one person by the time of the second follow-up interview and people they disclosed to usually supported their study participation. Men who were single disclosed mostly to relatives and friends, while married participants disclosed mainly to their partners, friends, and siblings.

Selective disclosure – For almost all trial participants, disclosing participation was specific and limited to certain individuals or categories of people. Participants feared compromising their confidentiality and privacy, reducing their worries about being seen to be taking part and to avoid any issues to do with stigma and social harms. They noted that they told only those that they felt mattered in the case of their safety, wellbeing or support.

“I can only tell that [disclose] to my person whom I trust. I can tell it to my parent or my sibling so that in case I find a problem, they can realize or understand that I once informed them that I was in the hospital (research centre) for this or that reason, and they can know what to do. But a person you meet on the way or a friend of yours whom you meet in eating and drinking places, if you tell him/her such information and you happen to suffer from some mild fever, they will say; ‘You see, we told you that what you had joined was no good. You are the one who has put yourself in the dangerous situation well aware of what you were doing’.” (female aged between 31 and over 35 years)

Unintended disclosure – There were reports of unintended disclosure whereby participants were compelled to reveal their participation due to suspicions raised by their regular travels to the clinic, PrEP pills (which were mistaken to be for HIV treatment), others seeing the information sheets, phone calls from study staff to contact persons, as well as rumours in the community. In most cases, this kind of disclosure resulted in challenges, raising the fears of harm and stigmatisation.

“…sometimes they would call her (the parent) if my number was off, but she was not aware of what was going on. She would just tell me that health workers in xx needed me. One day, after my clinic visit, I went home with my ‘bottles’ in the bag. I think she checked my bag and saw them; I found her almost sick and crying…” (male aged between 18 and 24 years)

In such situations, as in the example above, individuals would disclose their participation as well as provide information about the trial. Occasionally participants would provide HIV test results or, in the case of a sexual partner, go for couple HIV counselling and testing to prove they did not have HIV.

## Motivations for Disclosure

### Obtaining support and approval

Reasons for disclosing participation in the HIV vaccine trial varied and these usually ranged from self-focused goals like a desire to participate openly and comfortably free from the stress associated with concealment, to gaining approval and support from others, especially in terms of money for transport to the clinic and reminders about clinic appointments, swallowing study PrEP and filling in diary cards. Others disclosed because they wanted to keep their immediate family in the know, just in case they got any health complications, their people could know the possible cause.

Participants, especially the women, who had not disclosed their participation to their partners before building a more established relationship felt it was important to disclose after they moved in with/married them, since it would be hard to attend research visits, hide the pills or the diary cards. They felt that non-disclosure could result in non-adherence to the study requirements or social harm. For example, one woman had indicated at baseline that marriage was the only thing that could make her withdraw from the trial as she could not tell her partner about participation. Surprisingly, she got a stable partner by the second interview and disclosed to him about her participation in the trial while they were still courting. She noted that he continued loving her even after she disclosed and according to her, he understood and even provided support like money for transport to the clinic:

“By the time I married him, he already knew that I was part of a study, but he went ahead to court me up to marriage. Even though he did not know the type of drugs I was swallowing, when he saw them (the PrEP), it was easy for me to explain to him. After explaining, we went for couple HIV counselling and testing for him to confirm that I did not have HIV. In fact, I brought him here (to the study clinic) and the doctor explained it well to him.” (female aged between 18 and 24 years)

Although participants desired to disclose to the people they felt mattered in their lives or should know about their participation in the vaccine study, some lacked the confidence or the correct study information to provide to those they might disclose to, which in some cases prevented disclosure, as a failure to clearly explain the trial attracted or would attract negative reactions including disapproval. In other cases, the information they gave was distorted by other people who gave conflicting information thereby negatively influencing participants’ partners and, or families to the point of disapproving or blocking their participation. Among those who gave conflicting information were members of the community, those screened out of the study and, to a lesser extent, community health care providers. This happened especially during the initial stages of the study. Participants commonly reported how other people interfered with their participation by scaring their families and partners through giving misleading or incorrect information about HIV vaccines.

“At first, he used to ask me, what they were treating me for, I told him I was participating in a research study, but he would say I was being used as an experimental subject (guinea pig) and it will result in severe complications, and I end up lame because of those injections. I explained to him that they monitor us for up to three weeks after receiving the injection…It went on for some time but after that, whenever I took home medicine say for gonorrhoea, and tell him that I had got it from XX (the study clinic), he would also take some of it. Then he would give me lifts to the taxi stop whenever I was coming here.” (female aged between 18 and 24 years)

### Altruism and advocacy

Some participants disclosed their involvement because they felt a duty to contribute towards ending of HIV, and others viewed disclosure as an act of helping others by raising awareness of the available services in the trial and encourage or motivate other people to join and access these preventive services. They felt an obligation to help peers by informing them about the trial so that they could also join and benefit.

“There are some friends of mine whom I encouraged to join, and they joined. They were at a very high risk of getting HIV, yet they were not using any preventive measures.” (female aged between 18 and 24 years)

“I disclosed to my friends to motivate them to participate. When my friends ask why I am always travelling to XX, I tell them I am participating in research to help curb the spread of HIV.” (male aged between 25 and 30 years)

At times, participants viewed their involvement as a form of social responsibility whereby the trial outcomes could benefit not only them but also public health.

“I would wish to receive the vaccine and motivate others by serving as an example. Some of our people will only participate in anything after seeing others participate”. (male aged between 31 and 35 years)

So, this sense of altruism strengthened participants’ openness to discuss the trial with other people hoping that their disclosure might inspire trust in the trial and influence others to take part.

### Factors that facilitated Disclosure

The dissemination of information about the trial by study staff or fellow participants provided support to participants in countering rumours about the trial. In some cases, the study staff would talk to the family members to dispel misconceptions. Other participants who wanted to participate in the trial but feared disclosing for fear of not being believed by their partners, especially in the case of vaccine-induced sero-positivity (VISP) were also assisted by the trial staff to disclose to their partner or other family members.

“Remember my parent never went to school; I had to get someone to explain to her. I got someone who I gave the documents to read to her, but she broke into tears thinking her child is already HIV positive…” (male aged between 18 and 24 years)

“If you know that your relatives don’t know that you are here (in the trial), you are making a mistake. The doctors can talk to the partner and relatives if possible. That has ever happened to me.” (male FGD)

“My partner was aware that I am participating in the trial as the team (study staff) has ever contacted her through the phone trying to reach me, and she was okay with it. However, some people at one time scared her that I would get harm to the extent of failure to have children. I explained to her, and she understood.” (male aged between 18 and 24 years)

However, participants felt that disclosing participation was not always easy.

“I told my partner that I am participating in research on an HIV Vaccine to see if it can prevent HIV. Although the partner eventually understood; it was not an easy task, and I know many participants are going through the same difficulties. It is always hard if one of the partners is in the trial; it gets difficult to explain and convince the other especially the male partner to allow the female partner, for he would not be sure of the whereabouts of the female partner’’. (male aged between 31 and 35 years)

### Concerns about disclosing participation

Notwithstanding the fact that disclosure supported participation, there were major concerns about disclosure. These included a fear of being associated with or being labelled as having HIV or being promiscuous or unfaithful. For others, there was a fear of having to answer to a lot of questions which participants were not ready or able to, as they in some cases lacked the correct information. In cases where participants were suspected to have HIV, this would call for couple HIV counselling and testing, something that participants viewed as worrying because of vaccine induced sero-positivity (VISP):

“I have not disclosed to my partner because she might think I have HIV, yet I will not be able to take her for an HIV test due to VISP.” (male aged between 18 and 24 years)

For some women, they feared that their partners could stop them from participating if they disclosed, yet such women felt at risk of HIV and felt protected in the trial. Due to these reasons, participants feared to damage their relationships as it could result in losing financial support, especially for the women in case they broke up with their providing partners. However, non-disclosure of participation also came with its challenges as individuals who never disclosed not only failed to adhere to the trial requirements but kept lying about their movements, which resulted in mistrust, domestic violence and other forms of social harm.

He said, “I am healthy with no health issues. We both tested negative for AIDS (HIV) and saw each other’s results, now you are getting HIV vaccines, I am not ready for that.” That is how the man started changing and separated with me and changed his telephone numbers. I think they gave him more rumours about the study. He once called and said, “I loved you but lost interest when I heard about your participation in the study.” (female aged between 25 and 30 years)

Men, as well as women who had some form of independence and those who were transparent about their involvement in the vaccine trial participated more comfortably compared to those that never disclosed. It was interesting to note that some male participants whose partners did not trust their fidelity tended to support their participation in the study so that they could remain free from HIV.

Concerns about disclosing participation were exacerbated by the limited information that communities had about the trial and trial vaccines in general. Participants reported how their families and friends worried about them getting side effects including HIV sero-positivity, disabilities including physical defects and sexual dysfunction and sterility as well as death. The family members did not stop at worrying but discouraged them from taking part and threatened not to support the participants in case they experienced any of those side effects.

“Those people who were found with the HIV virus (study screen outs) are the ones who gossip about the trial. They say that the study staff injected me with the HIV virus. Some old person asked if I was aware of the drug that am being vaccinated with; I told him that I was sure it was safe depending on the study documents that I was given and how the trial is being managed. I even found a friend of mine who said that we should no longer be friends because of the vaccine that am being given, ‘I am scared of you’. People out in the community, whom the study screened out are the ones who say bad things about the study.’’ (female aged between 25 and 30 years)

“I do not want my close relatives to know about my participation in the trial because they would have very negative thoughts about my behaviour. I don’t want my parents to know it at all because they could think a lot about me”. (female aged between 25 and 30 years)

Participants highlighted the importance of engaging communities through sensitizing and providing them with information. Some suggested that some members of the community should be invited to visit the research centre and get an understanding of the study procedures and processes so that they get clear information of what to pass on to the rest.

### Benefits of disclosing participation

Disclosing participation in the trial in some cases facilitated people taking part: those who openly discussed their involvement in the trial benefited by eventually getting logistical and emotional support, especially from partners and family members. The support came up after these other people clearly understood the trial and how it would benefit participants and the community at large, in case the trial vaccines proved to be effective. Participants encountered less resistance or disapproval and even got support. This support came mainly in the form of transportation to clinic visits or financial assistance. Women whose partners had motorcycles or cars reported that they sometimes drove them to the clinic. Other family members also offered money for transport, while friends supported by lending money for transport to the clinic or taking care of the participants’ children or businesses while they travelled for their clinic visits. The other support was in form of encouragement and reminders to take their PrEP or about their clinic appointments.

“My parent is aware; sometimes, she provides me with transport money to enable me to get to the clinic…I had to tell her about the study to lessen her fears and good enough, I had the information sheet I had obtained from XX that guided me to explain the study very well. So, that information dismissed those fears that whoever was referred to XX the research centre has HIV. My parent supports me, but this came about after I had explained to her about the study, its purpose and future benefits to the entire community. Even when my clinic appointment dates reach, she usually reminds me to come for my visits”. (male aged between 25 and 30 years)

Participants’ narratives further revealed that individuals who disclosed their participation were able to participate freely without having to hide or to lie about their whereabouts or movements.

“…if your partner knows that you have gone to the clinic and you have explained the processes that you go through, I do not think it could be a problem. But if you leave home without his knowledge, for example, there are those who pretend to have gone to the garden, but they instead come to the clinic. Even the last time I was here, I was seated next to a woman and when her phone rang, she told me to tell the man on the phone that we were in the garden. That is what I told the man, but unfortunately, the man I was telling we were in the garden had followed his partner and was right there (at the clinic)”. (female aged between 25 and 30 years)

Looking at the findings through the lens of the DPM, we can see that when participants considered disclosure, they weighed up the potential benefits, such as social support, discovery of an effective vaccine, and increased trust in research, against risks, including stigma, discrimination, or misunderstanding from their communities. Those who perceived disclosure as leading to positive reinforcement and emotional support were more likely to remain engaged in the trial, while those who anticipated negative consequences might have declined participation, withdrew or never adhered to trial procedures.

## Discussion

This study set out to investigate how participants in an HIV vaccine and PrEP trial disclosed their taking part in the trial, and how this impacted on participant engagement. We found limited disclosure at baseline, which was a result of lack of prior knowledge about HIV trial vaccines, however, as participants gained more knowledge and confidence in the trial, disclosure of their taking part increased, leading to increased social support and improved trial participation, as well as a reduction in social harm.

The findings indicate that the limited knowledge of HIV trial vaccines among participants triggered scepticism and absolute denial of the vaccine’s existence when participants attempted to discuss it within their communities (19), since they could not clearly explain the trial. This lack of understanding about HIV vaccines was further compounded by misconceptions around vaccines functionality, likening it to direct exposure to the virus itself, relating it to their prior knowledge of how other routine vaccines worked (20). The misconceptions also hindered recruitment efforts, as participants who attempted to disclose or encourage others to participate faced resistance due to overwhelming doubts and questions from potential recruits, which questions participants could not clearly answer. There was a consensus among participants that community sensitization and education on how vaccines are developed and tested would be crucial in fostering confidence in the vaccine trials and consequently enable trial participants discuss them freely and comfortably.

Participants disclosed their involvement in the trial to varying degrees, depending on their relationships and perceived consequences of disclosing. Disclosure was more common to people whom participants felt mattered, especially when logistical support or approval was needed for clinic visits. Individuals who disclosed taking part tended to exhibit improved participation in terms of honouring research visits and adhering to trial procedures. This finding is in agreement with findings from other studies, whereby participants who discussed their participation in health interventions with key people in their lives adhered better and received social support thus displaying better prevention outcomes (21–25).

In our study, disclosing participation not only engendered social support, but created awareness as well as demystifying the misconceptions and clearing misinformation about the trial within the communities. Other studies have had similar findings of how disclosing yields benefits such as greater social support and increased health outcomes (21, 26).

However, disclosure was not always straightforward and while disclosing was in most cases associated with the desire to traverse the concerns/barriers that were foreseen and hindrances to participation or adherence to the trial processes, some participants chose not to disclose due to fears of stigma, misinterpretation, and community gossip (25). The stigma associated with HIV-related research meant that participants risked being labelled as having HIV, being promiscuous, or unfaithful. This was particularly problematic for women, as disclosing participation could lead to relationship conflicts, loss of financial support, or even domestic violence (27–29). This calls for widespread education on HIV vaccines to counter misinformation and reduce stigma.

In addition, this stigma from HIV-related research can cause individuals to struggle with making decisions on whether to disclose, how, when and whom to tell, since some disclosures may be related to more negative outcomes if not well calculated, rendering concealment a more reliable prospect of one’s wellbeing than when a person discloses (30). This could be one of the reasons why some participants in our study tended not to disclose, and even those who disclosed did it with particular categories of people and not others. They also calculated how much information to give out to the different categories of people. Other studies have indicated this selective disclosure (23–25). However, secrecy or issues around non-disclosure can lead to undesirable outcomes like non-uptake or non-adherence to study procedures, as well as social harm, as observed with this study. This calls for a more nuanced understanding of the complex nature of disclosure within and under different circumstances. Interventions to empower people to reach decisions about disclosing, especially in cases of stigmatizing conditions are important. An example is the Honest, Open, Proud (HOP) program, which supports disclosure decisions of mental health problems and other stigmatized conditions (31)

Since lack of clear information regarding the trial and its components was a barrier to disclosure, trial implementers need to always ensure that trial participants clearly understand the trial. This is important for participants’ consumption and for how they disseminate information about the study. This could be done through repeated information giving and assessment of understanding over the duration of the trial.

A critical insight from our findings is that individuals who had participated in vaccine trials before were considered as trusted sources of information. The allusion that hearing directly from those who had completed the vaccination schedule without experiencing complications would encourage others to participate was evident when individuals who were hesitant to take part, decided to participate after witnessing others safely participating in the trial (32–34). In view of this, utilization of former trial participants as advocates could increase trust and credibility in vaccine trials.

The findings also indicate that assisted disclosure by study staff is crucial in easing concerns, especially with participants who are not able to disseminate the trial information to others. Participants eventually got approval and support after they brought their partners to the research clinic for more information about the trial.

Community perceptions influenced the experiences of trial participants considerably. Initial reactions to the vaccine trials were largely negative, driven by misinformation and fear.

Rumours about participants being injected with HIV or suffering severe health complications discouraged many from joining the study. Some participants even faced social exclusion, with friends and family distancing or threatening to exclude them. However, with time, as participants gained more knowledge about the vaccine and its safety, they were able to counteract some of these misconceptions, reducing resistance from their social circles. Many suggested that proactive community engagement, including outreach programs and inviting community members to observe study procedures first-hand, would help dispel fears and encourage informed participation. Our findings illustrate how community norms and attitudes toward HIV research shape disclosure behaviours, which could ultimately affect recruitment, retention, and adherence to trials.

The strength of this study lies in its longitudinal nature, which provided insights into changes in participants’ experiences with disclosure and adherence behaviours over the different phases of the trial. Also, the selection of participants based on various characteristics ensured a diverse set of perspectives, enhancing the study’s validity, while the data from the different sources (IDIs, FGDs) enhanced the credibility of the data in addition to allowing for a deep exploration of participants’ experiences, allowing the study team to identify concerns and enabling trial staff to address them in real time resulting in improved uptake and conduct of the trial.

A limitation to the study was the limited generalizability that besides this being just a small number of the trial participants, which may limit the ability to capture the full range of experiences, the study focused on a specific population of individuals at high risk of HIV, recruited from areas of high HIV prevalence and the findings from their data may not be generalizable to other people in the general population. Additionally, despite the study being part of a larger multi-site trial, these are findings from a single site in Uganda, and this may limit the applicability of the findings to other trial sites where contextual factors may differ.

### Conclusion

The findings highlight the significant knowledge gaps, fears, and societal influences that impact participation in HIV vaccine trials. While disclosure had both benefits and challenges, it was evident that informed communities were more likely to support trial participation. Moving forward, targeted educational/awareness campaigns and community engagement strategies will be essential in fostering a supportive environment for HIV vaccine research and development through increased participation, strengthening retention and adherence, and reduction in social harm.

## Data Availability

Illustrative data supporting the study’s findings are presented in the manuscript. Anonymized transcripts are not publicly available, as they may contain information that could compromise participant privacy.

## Acknowledgements

We acknowledge the contributions of the PrEPVacc study team and all the participants. Special thanks to the MRC/UVRI & LSHTM Uganda Research Unit for all their contribution and support. We thank the Community Advisory Board (CAB) members who worked as a link and supported this research within the communities.

## Authors’ contributions

Conceptualisation of the study: RK, SK, ER, JS; investigation: ER, JS; methodology: SN, RK, JS; data curation, visualisation and validation: SN, RK, DK, GN; formal analysis: SN, RK, DK, GN; supervision: RK, JS; writing - original draft, writing: SN; writing – review and editing the manuscript: SN, RK, DK, GN, SK, ER, JS. All authors reviewed and approved the final version of the manuscript.

## PrEPVacc Team (Uganda site)

MRC/UVRI and LSHTM Uganda Research Unit, Masaka, Uganda (Site):

Pontiano Kaleebu, Eugene Ruzagira, Freddie Mukasa Kibengo, Janet Seeley, Sylvia Kusemererwa, Martin Onyango, Yofesi Nikweri, Anta Kabarambi, Ubaldo Bahemuka, Jonathan Kitonsa, Shamim Ssendagire, Masawi Sylvia, Vincent Basajja, Beatrice Kimono Washi, Elizabeth Mbabazi, Cissy Lillian Nalubega, Margaret, Lydia Wesonga, Sarah Nakato, Racheal Wanyana, Hadijah Naluyinda, Coleman Tayebwa, Philip Kibuuka, Mary Namukisa, Grace Muyingo, Ivan Kisubika, Doreen Asio, Angel Nansere, Ssemaganda Henry, Victoria Mugwaneza, Safinah Katana, Solomon Wasswa, Mande Sulait, Kitumba Joseph, Josephine Bayigga, Ventus Kwigenga, Hussein Nyombi, Ronald Kiwanuka, Baker Ssebunya, Mike Mukasa, Sarah Nakamanya, Rachel Kawuma, Dennis Kibuuka, Edith Nalwadda, Abdmagidu Menya, Ayoub Kakande, Shamim Nabukenya, Paddy Kafeero L, Sandra Nabalayo, Joanita Nassali, Justin Okello, Gertrude Mutonyi, Namirembe Aeron, Naphtali Erima, Victoria Nakibirango, Fulgensio Bbosa, Barbara Babirye, Julius Mutagubya, Nakkazi Sophia Kakyama, Shanila Nasejje, Penelope Akankunda, Dianah Tuwangye, Alex Mutazindwa, Florence Nambaziira Muzaale, Edward Muhigirwa, Didas Mushabe, Tobias Vudriko, Flavia Kisakye, Peter Ejou, Zephyrian Kamushaaga, Jenifer Serwanga, Betty Oliver Auma, Susan Mugaba, Angela Namuyanja, Regina Nanyunja, Solomon Opio, Ben Gombe, Godfrey Matovu, Joan Bwandinga, Nasim Kyakuwa, Willy Fred Ochola, Faith Wamalugu, Esther Nabanoba, Juliet Bukenya, Charline Katurole, Priscilla Balungi, Dora Jocelyn Mulondo, Bruno Mukundane, Joyce Nabunnya, Nsimire Juliet Sendagala, Ayebazibwe Gloria Kakoba, John Vianney Kagaba, Kitonsa Mugagga Derrick, Fiona Nangobi, Wilson Kakeeto, Paul Taire, Bridget Muhofa, Mugagga Kyeyune, Phiona Nabaggala, Dorothy Abigaba, Bernadette Nayiga Kalanzi, Sheila Kansiime, Christian Holm Hansen, Fr Emmanuel Katabazi (CAB representative).

## Trial oversight and management support

Jonathan Weber, Cherry Kingsley, Tom Miller, Sheena McCormack, Angela Crook, David Dunn, Henry Bern, Aminata Sy, Simona Salomone, Liz Brodnicki, Sarah Joseph, Claire Wenden, Giuseppe Pantaleo, Song Ding, Charlotta Nilsson, Arne Kroidl, Julie Fox, Gustavo Doncel, Allison Matthews, Jim Rooney, Carter Lee, Merlin Robb

## Monitoring

IAVI: Kundai Chinyenze, Jacqueline Musau, Mabela Matsoso, Mary Amondi, Ansuya Naidoo, Paramesh Chetty, Anne Gumbe

